# Impact of Artificial Intelligence Assistance on Efficiency, Ergonomics, and Measurement Consistency of Abdominal Ultrasound

**DOI:** 10.64898/2026.11.23.26368613

**Authors:** Richard G. Barr, Jing Gao, Adrian Lim, Brandy Neill

**Affiliations:** Northeastern Ohio Medical University, Rootstown, Ohio; Southwoods Imaging, Youngstown, Ohio; Rocky Vista University, Montana College of Osteopathic Medicine, Billings, Montana; Department of Imaging, Imperial College London and Healthcare NHS Trust, London, UK

**Keywords:** abdominal ultrasound, artificial intelligence, ergonomics, musculoskeletal disorders, sonography, workflow efficiency

## Abstract

**Objectives:** Evaluate how the addition of AI-assisted image acquisition and measurements affects the efficiency, ergonomics, accuracy, and consistency of abdominal ultrasound examination.

**Methods:** Randomized, crossover study utilizing an abdominal ultrasound system with integrated AI features. Experienced sonographers (N=33) performed standardized examinations with and without AI-assisted features enabled. Metrics of timing and control interactions by the non-scanning hand were acquired. Movements of the non-scanning hand were recorded during a subset of examinations (N=17) to evaluate distance traveled with and without AI. An offline measurement exercise (N=32) using standardized image datasets assessed agreement between AI-generated and manual measurements and the effect of AI assistance on the consistency of measurements across users.

**Results:** AI assistance reduced mean exam time by 2.23 ± 0.44 minutes (95% CI: −3.10, −1.36; *P*<0.001); independent of participants’ age, experience, or the sequence of AI-assisted vs manual scans. AI assistance reduced mean distance traveled by the non-scanning hand by 49.2% (*P*<0.001). AI assistance also reduced overall control interactions by the non-scanning hand by 36%, touchscreen reaches by 41.5%, interactions with the control panel by 23.4%, and mouse movements and clicks by 54.2% and 61.1%, respectively (*P*<0.001 for all). During offline measurement, users accepted 94.3% of AI-generated measures without modification. AI assistance reduced inter-user measurement variability by 74%.

**Conclusions:** AI assistance significantly reduced exam time and improved ergonomics compared with manual scanning and supported more standardized measurements across users. These findings support the integration of AI-assisted tools into imaging workflows to enhance workflow efficiency, ergonomics, and diagnostic consistency.

## INTRODUCTION

Ultrasound is a fast-growing imaging modality that plays a critical role in screening, diagnosis, surveillance, and interventional guidance across a broad range of specialties and clinical settings.^1,2^ Abdominal ultrasound is one of the most frequently requested investigations, accounting for 60% or more of general diagnostic ultrasound examinations.^3–5^

Recent evidence indicates that demand for sonography is increasing faster than the supply of trained sonographers. In the United States, the number of ultrasound examinations performed in outpatient settings increased by 55.1% between 2011 and 2021. Over the same period, the number of sonographers increased by just 43.6%.^2^ This deficit places a considerable burden on individual sonographers, some of whom conduct more than 15 examinations per 8-hour workday.^6,7^ In addition, as scan protocols become more comprehensive and the demand for advanced measurements grows, the duration and physical intensity of examinations has increased—placing significant ergonomic stress on sonographers.^8^

Work-related musculoskeletal disorders (WRMSDs) are a persistent occupational hazard in sonography. Surveys indicate that between 80%–90% of sonographers experience scanning-related pain, with a significant proportion performing scans while in pain.^6,9^ A 2024 systematic review and meta-analysis encompassing 30 studies and nearly 14,000 sonographers estimated the overall prevalence of musculoskeletal disorders among sonographers at 75.8%.^10^ Similarly, a U.S./Canadian survey of more than 3500 ultrasound users found that 86% regularly experienced musculoskeletal discomfort directly attributable to their work, most often in the shoulder, neck, and wrist/hand.^11^ In a U.S. survey of nearly 8,000 credentialed sonographers, 46.3% of those whose primary specialty was abdominal ultrasound reported work-related musculoskeletal injuries and 64.5% reported scanning in pain.^7^

The consistency of these findings establishes WRMSDs as a significant and enduring occupational health problem in sonography, with wide-ranging impacts on the quality and consistency of sonographic examinations. WRMSD-associated discomfort has been linked to an estimated 5%–7% decline in productivity and work ability and may also contribute to workforce attrition.^8,12,13^ Indeed, WRMSDs are potentially career-ending for approximately one in five affected sonographers.^8,10,14^

Although discussions of the ergonomics of ultrasound often emphasize the probe hand, the non-scanning hand is also exposed to repetitive and sustained mechanical loading. Operating the control panel, touchscreen, knobs, calipers, and trackball while simultaneously maintaining probe contact creates a bilateral task in which one arm is subjected predominantly to static force while the other is exposed to reaching and repetitive user-interface interaction.^15^ To prevent WRMSD, industry standards set by the Society of Diagnostic Medical Sonography stress the importance of minimizing non-neutral and static postures, and of avoiding excess reach and abduction of the non-scanning arm.^16^

Technological solutions that reduce repetitive motion, cumulative strain, and exam time are essential to addressing the clinical and occupational health needs of resource-constrained ultrasound labs. To this end, artificial intelligence (AI) technologies are increasingly being investigated as a means to automating labor-intensive tasks and alleviating physical strain. AI tools such as automated imaging protocols, real-time anatomical labeling, measurement acquisition, and organ segmentation have the potential to optimize clinical performance while reducing the cumulative physical burden on the sonographer.^17–19^

The purpose of this study is to determine if AI-assisted image acquisition and measurements can improve the efficiency, ergonomics, and consistency of abdominal ultrasound examination.

## MATERIALS AND METHODS

The study sponsor (Siemens Healthineers) contracted with AIUM to recruit sonographers to participate in the study. Participants were selected from a list of volunteers provided by AIUM. Participant selection focused on individuals with experience in abdominal ultrasound and sought to ensure diversity in credentials, education, age, and work environment. Participants were not informed of the study sponsor, study hypotheses, or the commercial name of the AI feature before participation, and provided informed consent via a standardized, IRB-approved consent form. All participants were new users of the AI feature, AI Abdomen, and only one participant reported prior experience on a Siemens ultrasound system.

### Study Design

This study utilized a two-period crossover design in which participants allocated to perform abdominal ultrasounds on volunteer models, either beginning with AI assistance and progressing to a manual evaluation or beginning with manual acquisition and progressing to AI assistance using an alternating assignment procedure based on arrival order the day of the study. Successive participants were assigned to opposite sequences to maintain balance between treatment orders. Sequence (AI then Manual or Manual then AI) was subsequently included as a factor in the statistical analysis to evaluate potential order effects. Each participant was provided with a 10-minute break between scans. A standard imaging protocol was followed for both the AI-assisted and manual examinations, as delineated in **Table 1**. Participating sonographers served as their own control to minimize inter-individual variability.

**Table 1.** Artificial Intelligence (AI)-Assisted and Manual Protocols.

| **AI Protocol** |  |  | **Manual Protocol** |  |
|---|---|---|---|---|
| **Aorta Sagittal**<br>• 2D image of aorta long (mid-distal area)<br>• Freeze | Image capture |  | **Aorta Sagittal**<br>• 2D image of aorta long (mid-distal area)<br>• Freeze | Annotate image (touchscreen or typing) / Image capture |
| **Inferior Vena Cava (IVC) Sagittal**<br>• 2D image of IVC Long (Prox area)<br>• Freeze | Image capture |  | **Inferior Vena Cava (IVC) Sagittal**<br>• 2D image of IVC Long (Prox area)<br>• Freeze | Annotate image (touchscreen or typing) / Image capture |
| **Pancreas Transverse**<br>• 2D image of pancreas transverse<br>• Freeze | Image capture |  | **Pancreas Transverse**<br>• 2D image of pancreas transverse<br>• Freeze | Annotate image (touchscreen or typing) / Image capture |
| **Pancreas Sagittal**<br>• 2D image of pancreas at the head<br>• Freeze | Image capture |  | **Pancreas Sagittal**<br>• 2D image of pancreas at the head<br>• Freeze | Annotate image (touchscreen or typing) / Image capture |
| **Left Liver Sagittal**<br>• 2D image of left liver sagittal medial area (no aorta/IVC)<br>• Freeze | Image capture |  | **Left Liver Sagittal**<br>• 2D image of left liver sagittal medial area (no aorta/IVC)<br>• Freeze | Annotate image (touchscreen or typing) / Image capture |
| **Left Liver Transverse**<br>• 2D image of left liver transverse at level of left hepatic vein or left portal vein<br>• Freeze | Image capture |  | **Left Liver Transverse**<br>• 2D image of left liver transverse at level of left hepatic vein or left portal vein<br>• Freeze | Annotate image (touchscreen or typing) / Image capture |
| **Right Liver Transverse**<br>• 2D image of right liver transverse at level of right hepatic vein or right portal vein<br>• Freeze | Image capture |  | **Right Liver Transverse**<br>• 2D image of right liver transverse at level of right hepatic vein or right portal vein<br>• Freeze | Annotate image (touchscreen or typing) / Image capture |
| **Right Liver Sagittal**<br>• 2D image of right lobe of liver (at the level you would take the liver length)<br>• Freeze | AI measurement / Image capture |  | **Right Liver Sagittal**<br>• 2D image of right lobe of liver (at the level you would take the liver length)<br>• Freeze | Annotate image (touchscreen or typing) / Activate calipers / Select measurement label [liver length] / Measure liver length / Image capture |
| **Common Bile Duct (CBD)**<br>• 2D Image of CBD at level of biliary tree or pancreatic head<br>• Activate color if needed<br>• Freeze | AI measurement / Image capture |  | **Common Bile Duct (CBD)**<br>• 2D Image of CBD at level of biliary tree or pancreatic head<br>• Activate color if needed<br>• Freeze | Annotate image (touchscreen or typing) / Activate calipers / Select measurement label [CBD] / Measure CBD diameter / Image capture |
| **Gallbladder Sagittal**<br>• 2D image of gallbladder sagittal (may decrease sector width and adjust as needed)<br>• Freeze | Image capture |  | **Gallbladder Sagittal**<br>• Decrease sector width<br>• 2D image of gallbladder sagittal<br>• Freeze | Annotate image (touchscreen or typing) / Image capture |
| **Gallbladder Transverse**<br>• 2D image of gallbladder transverse (may decrease sector width and adjust as needed)<br>• Freeze | AI measurement of gallbladder wall / Image capture |  | **Gallbladder Transverse**<br>• 2D image of gallbladder transverse<br>• Freeze | Annotate image (touchscreen or typing) / Activate calipers / Select measurement label [gallbladder wall thickness] / Measure gallbladder wall thickness/ Image capture |
| **Right Kidney Sagittal**<br>• 2D image of right kidney sagittal<br>• Freeze | AI measurement of right kidney length / Image capture |  | **Right Kidney Sagittal**<br>• 2D image of right kidney sagittal<br>• Freeze | Annotate image (touchscreen or typing) / Activate calipers / Select measurement label [right kidney length] / Measure right kidney length / Image capture |
| **Right Kidney Transverse**<br>• 2D image of right kidney transverse mid-pole at level of the hilum<br>• Freeze | AI measurement of right kidney anteroposterior and width / Image capture |  | **Right Kidney Transverse**<br>• 2D image of right kidney transverse mid-pole at level of the hilum<br>• Freeze | Annotate image (touchscreen or typing) / Activate calipers / Select measurement label [right kidney anteroposterior] / Measure right kidney anteroposterior / Image capture |
| **Spleen Sagittal**<br>• 2D image of spleen sagittal<br>• Freeze | AI measurement of spleen length / Image capture |  | **Spleen Sagittal**<br>• 2D image of spleen sagittal<br>• Freeze | Annotate image (touchscreen or typing) / Activate calipers / Select measurement label [spleen length] / Measure spleen length / Image capture |
| **Spleen Transverse**<br>• 2D image of spleen transverse<br>• Freeze | AI measurement of spleen anteroposterior and width / Image capture |  | **Spleen Transverse**<br>• 2D image of spleen transverse<br>• Freeze | Annotate image (touchscreen or typing) / Activate calipers / Select measurement label [spleen anteroposterior] / Measure spleen anteroposterior / Select measurement label [spleen width] / Measure spleen width / Image capture |
| **Left Kidney Sagittal**<br>• 2D image of left kidney sagittal<br>• Freeze | AI measurement of left kidney length / Image capture |  | **Left Kidney Sagittal**<br>• 2D image of left kidney sagittal<br>• Freeze | Annotate image (touchscreen or typing) / Activate calipers / Select measurement label [left kidney length] / Measure left kidney length / Image capture |
| **Left Kidney Transverse**<br>• 2D image of left kidney transverse mid-pole at level of the hilum<br>• Freeze | AI measurement of left kidney anteroposterior and width / Image capture |  | **Left Kidney Transverse**<br>• 2D image of left kidney transverse mid-pole at level of the hilum<br>• Freeze | Annotate image (touchscreen or typing) / Activate calipers / Change from right to left on upper right / Select measurement label [left kidney anteroposterior] / Measure left kidney anteroposterior / Select measurement label [left kidney width] / Measure left kidney width / Image capture |
| **Report** | Image capture |  | **Report** | Image capture |
| **End Exam** |  |  | **End Exam** |  |

*Note*. 2D = two-dimensional; AI = artificial intelligence; CBD = common bile duct; IVC = inferior vena cava.

After completing the live scanning portion of the study, participants engaged in an offline measurement exercise using a custom software application on laptop computers. Participants completed measurements of standardized abdominal ultrasound images both manually and with AI assistance. During the AI-assisted segment, the system generated initial caliper positions which participants could accept without modification or adjust before saving the measurement. The sequence of manual vs AI-assisted measurement was randomly assigned.

The study design and informed consent forms were reviewed and approved by the sponsor’s institutional review board (WIRB-Copernicus Group IRB, Puyallup, Washington).

The study was conducted during the 2025 American Institute of Ultrasound in Medicine (AIUM) Annual Convention in Orlando, Florida. Nine ultrasound stations were installed in an exhibit hall; each contained an ultrasound system, patient bed, table, two chairs, a trash can, gel, electrodes, paper towels, and gowns (see **Figure 1**).

**Figure 1.**
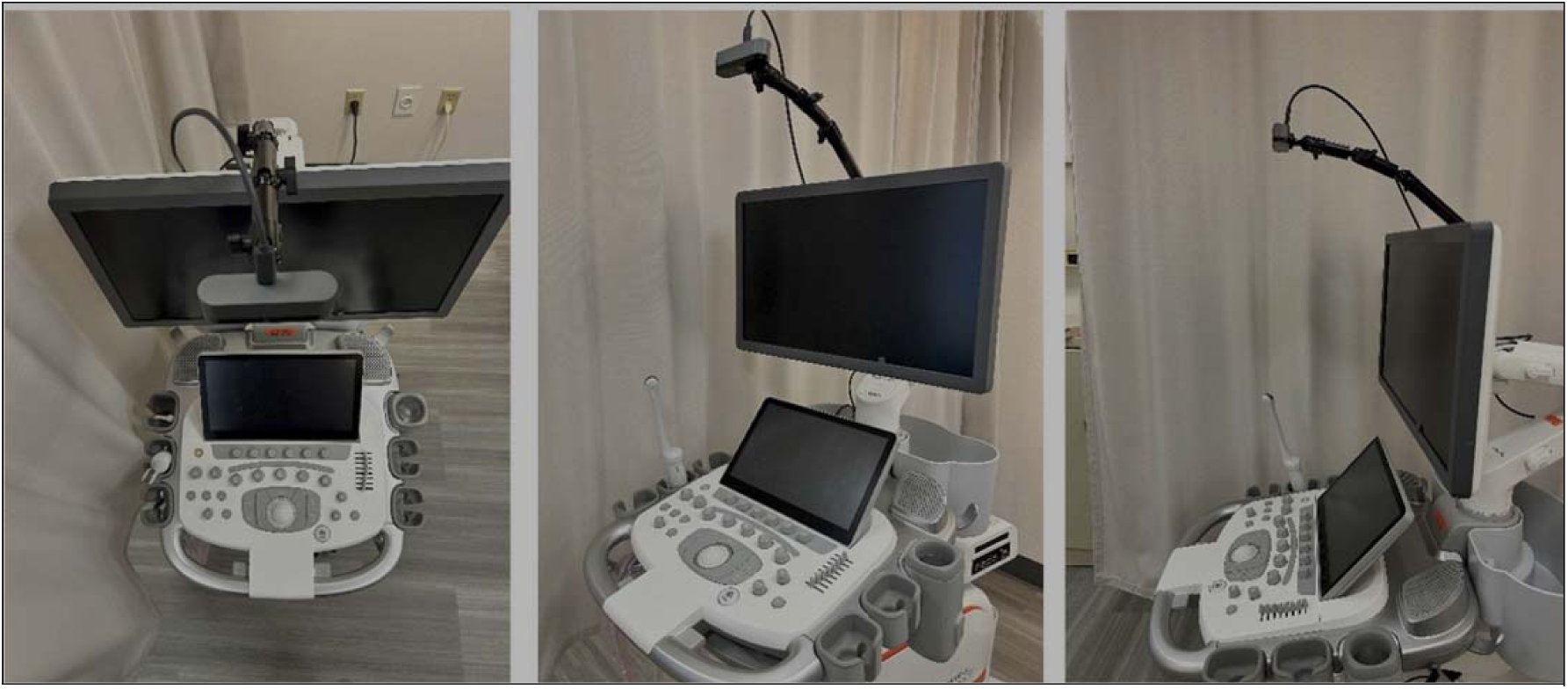
Scanning stations and hand tracking systems. Nine ultrasound stations were installed in an exhibit hall at the 2025 American Institute of Ultrasound in Medicine (AIUM) Annual Convention in Orlando, Florida. Four stations were outfitted with hand-tracking systems to record hand movements from an overhead perspective.

Participants received a copy of the protocol prior to the event and were given a 15-minute on-site introduction to the system, consisting of review of the control panel; an explanation of the touchscreen and how it presents functions based on image, view, and modality; instructions on manual measurements and capture for reporting; and how to enable AI. Participants were asked to concentrate on image acquisition and measurements and to minimize questions about features beyond those needed for the study tasks. Proctors were provided for each participant session. The proctors provided system introduction and overview of the scan protocol during the 15-minute introduction. After the exam period started, the proctors were instructed to only enforce that the scan protocol was followed and avoid providing guidance on button locations. Additionally, coaching on system operation would most likely only be requested during the first exam period. This would be included in the analysis of period to period differences where there were significant effects. To assess the impact of AI assistance on efficiency and ergonomics, high-definition (30 frames per second) video cameras were installed above four of the nine ultrasound stations to record the non-scanning hand’s interaction with the control panel. Cameras had a clear view of the touchscreen and physical controls operated by the non-scanning hand. For tracking purposes, sonographers wore a disposable examination glove marked with a circular sticker on the dorsum of the middle finger, as shown in Figure 2. Video footage was processed using Adobe After Effects (version 24.6.4), utilizing integrated tracking tools to measure motion from the start of image acquisition to the final image.

**Figure 2.**
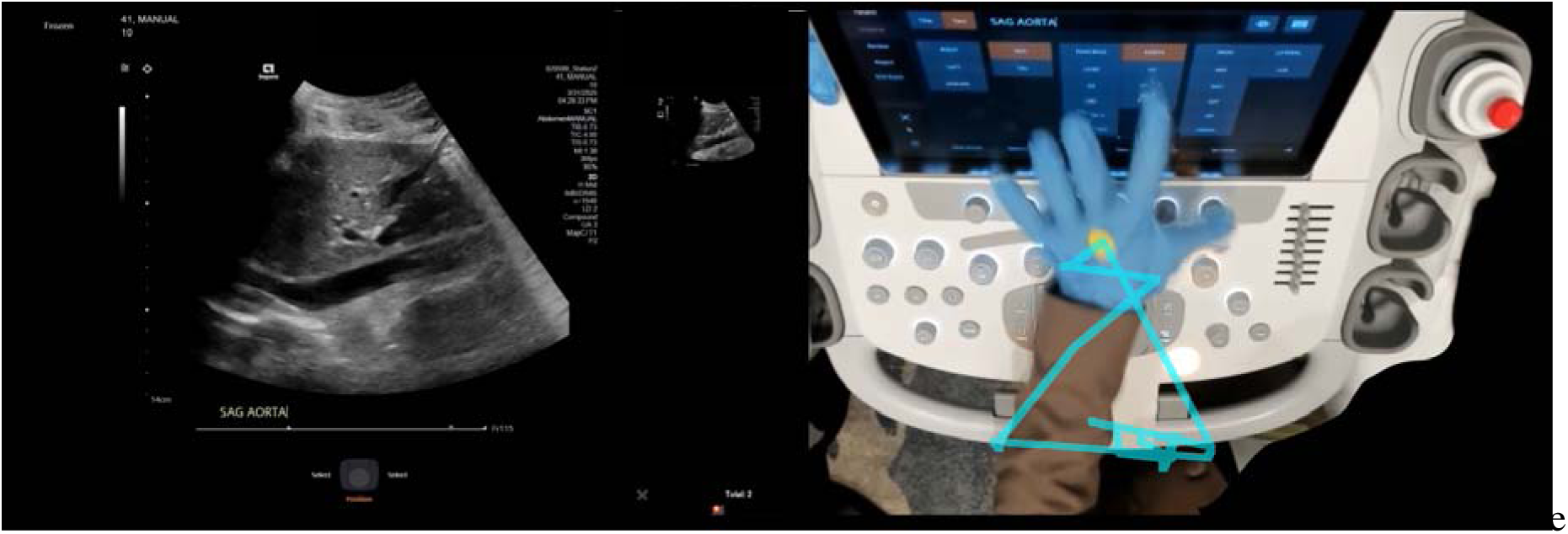
Hand tracking. Hand tracking for a manual scan, illustrating gloved hand with circular sticker on the dorsum of middle finger to assess hand movement over the control panel in correlation to the scanning image on the left.

### Study Participants

Ultrasounds were performed on 17 imaging models who had been pre-screened to confirm normal body habitus of less than 25 body mass index (BMI) and an absence of abdominal abnormalities. Models were recruited through a modeling agency and were compensated for their time through the agency; all provided informed consent via a standardized IRB-approved consent form.

### Abdominal Ultrasound System

The study utilized the ACUSON Sequoia ultrasound system (Siemens Healthineers, Issaquah, Washington), with integrated AI-assisted features (AI Abdomen) that received FDA clearance in 2024 (see **Appendix 1**). AI Abdomen automatically recognizes and annotates 17 anatomical views for a standard abdominal examination and performs 12 AI-assisted measurements allowing edits as needed. Laptop computers were used for an offline measurement exercise to test the consistency of measurements with AI assistance vs manual measurements.

### Outcomes

The primary endpoint was mean difference in examination time (in minutes) between manual and AI-assisted abdominal ultrasound exams. Secondary endpoints included ergonomic efficiency, ease of use and user satisfaction, measurement acceptance, and inter-user variability of measures obtained using AI.

Ergonomic efficiency was measured by comparing the average hand distance traveled (in meters) with and without AI assistance during a subset of examinations. In addition, interactions with system inputs by the non-scanning hand were assessed by counting the clicks on system controls, categorized according to principles derived from the Rapid Upper Limb Assessment (RULA) framework, as illustrated in **Figure 3**.^20^ Click interactions were grouped by specific input type: Touchscreen (press of touchscreen and soft key knobs below touchscreen); Control Panel (interaction with control panel); Mouse Move (movement of trackball); and Mouse Click (press of left or right mouse buttons).Ease of use and user satisfaction were assessed via a 16-item post-study participant survey (see **Appendix 2**)

**Figure 3.**
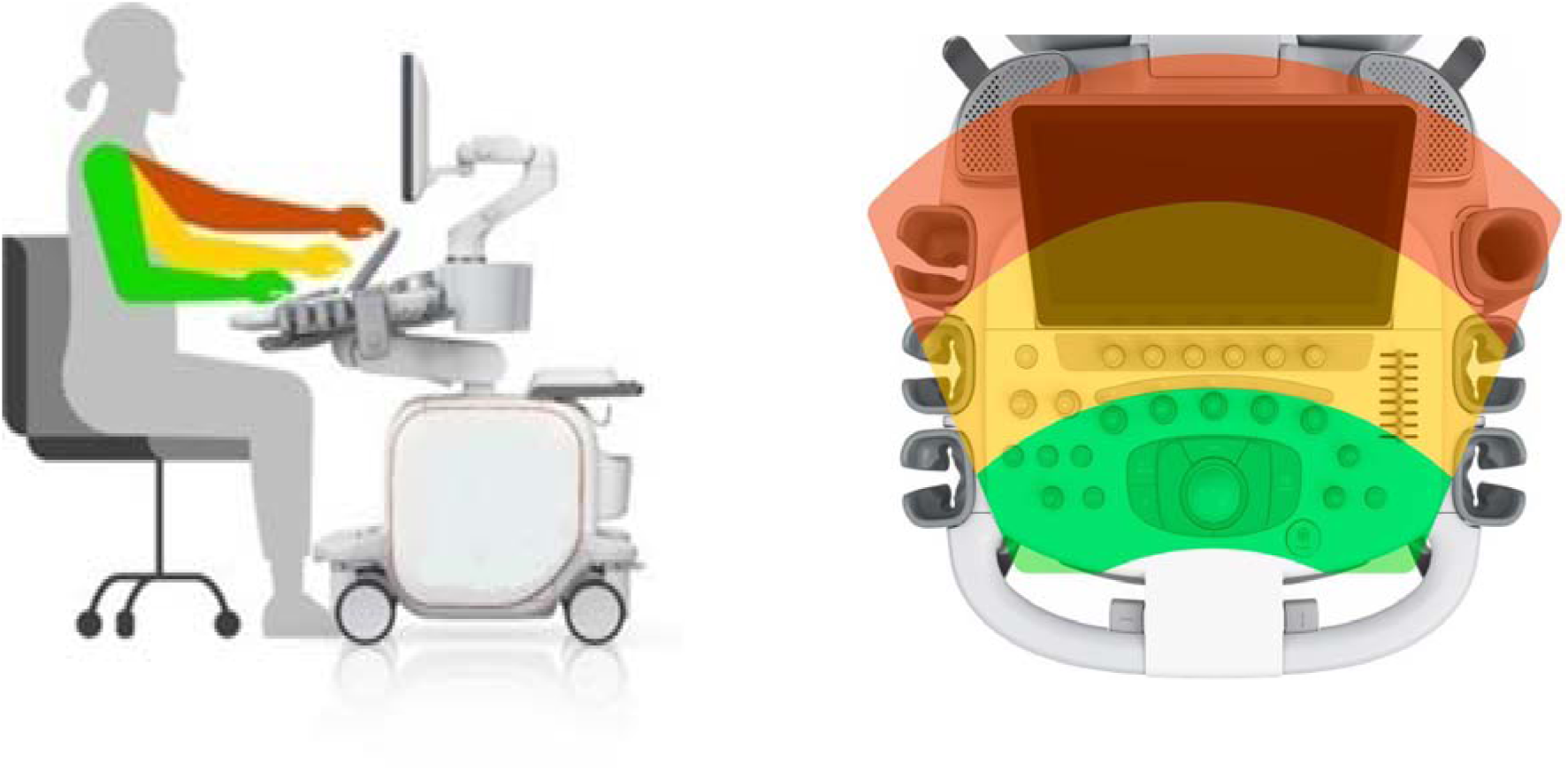
Ergonomic Reach Zones. The neutral (green) zone is recommended for all tasks, the secondary (yellow) zone is acceptable for occasional tasks, and the tertiary (red zone) is for infrequent tasks.

Measurement acceptance and inter-user variability were assessed by comparison of measurements obtained with and without AI assistance during an offline measurement exercise. This analysis was designed to assess agreement between AI-generated and manual measurements, quantify user acceptance of AI-generated measurements, and evaluate the effect of AI assistance on the consistency of measurements across users.

### Statistical Methods

The within-subject design of this study enabled direct comparison of key performance indicators, including total exam time, distance traveled by the non-scanning hand, and overall click count (number of button/knob touches per exam). Total exam time served as a surrogate for examination efficiency. Hand movement and click counts served as a surrogate for ergonomic burden. Sample size was selected as a minimum of 30 as a pragmatic size for a 2X2 cross-over design given the constraints of available systems, support staff, and participant availability during the AIUM conference. Over-recruiting was performed to ensure a minimum of 30 was achieved. For all analyses other than the offline measurements, the models were fit using restricted maximum likelihood estimation (REML). Statistical significance was evaluated using Satterthwaite’s approximation for degrees of freedom and defined as *P* < 0.05. To asses potential order effects inherent to the crossover design, Sequence (AI/Manual versus Manual/AI) and period (first period or second period in the crossover design) were included as fixed effects in the analysis. Statistical analyses were performed in R version 4.6.1 using the packages lme4 (v2.0.6), lmerTest (v3.2.1), emmeans (v2.0.4), car (v3.1.5), glmmTMB (v1.1.14), DHARMa (v0.5.0), tidyr (v1.3.2), dplyr (v1.2.1), and ggplot2(v4.0.3).

#### Exam Time Analysis

Because the experience of conducting the initial scan might have a positive influence on performance during the second scan, the magnitude of carryover effect was assessed for all comparisons. Thereafter, the effect of Sequence (AI/Manual versus Manual/AI), Period (first period or second period in the crossover design), and Method (Manual or AI) on examination time was evaluated using a linear mixed effects model with Participant fitted as a random effect.

#### Hand Distance Analysis and Ergonomics

Analysis of hand distance was conducted in a subset of examinations (n = 17) with Sequence, Period, and Method fitted as fixed categorical effects looking at distance traveled in meters by the non-scanning hand during each examination.

Interactions with system inputs by the non-scanning hand were assessed by counting the clicks on system controls across the four input devices. A linear mixed effects model was fitted for these data using participant as a random effect to account for repeated observations and participant-by-input-type as a random effect to account for differences in individual interaction styles. Fixed effects included Method, Period, and Sequence. Post hoc comparisons between input types were conducted using estimated marginal means with Holm-adjusted *P* values to identify differences by input between the Methods.

Because click counting can be associated with non-normal and sometimes skewed data, the linear mixed effects model was compared with a negative-binomial mixed model using likelihood-ratio tests, Akaike Information Criterion and Bayesian Information Criterion as a robustness check. The linear mixed effects model was found to be a better fit to the observed data.

The potential for carryover effects from the first period influencing the second period were also assessed through the Sequence factor. In the two-period crossover design, a significant effect for Sequence would be indicative of an order effect. A non-significant sequence effect was considered evidence that treatment order did not materially influence exam time.

#### Offline Measurement Analysis

For AI-assisted measurements, edit frequency and edit magnitude were evaluated. AI-generated measurements were considered accepted without modification when no caliper adjustment was made. Edited measurements were classified by the extent of user adjustment. Major adjustments were defined as combined caliper movement of at least 5 pixels and a resulting change in measured distance of at least 2% relative to the original AI-generated measurement.

Use of the same images for all participants eliminated acquisition variability and allowed direct assessment of inter-user measurement consistency. Measurement consistency was evaluated using the coefficient of variation (CV), calculated for each measurement type as the ratio of the standard deviation to the mean measured distance across participants. CV values were summarized across the offline measurement dataset for Manual measurements and for AI-assisted measurements after user edits, with lower CV values indicating greater inter-user consistency.

## RESULTS

### Participants

Demographics of the 33 volunteer participants who completed the two-examination protocol are presented in **Table 2** (one participant was excluded from the offline measurement analysis due to completion of only the Manual measurement portion of the task). Most had > 10 years in the field, but the group included some with < 5 years’ experience (6.1%) and some with > 20 years (42.4%). The majority (66.7%) had a bachelor’s degree or higher and 90.1% were registered diagnostic medical sonographers. Participants were predominantly right-handed scanners (84.8%).

**Table 2.** Sonographer Demographics.

| **Variable** | **N (%)** |
|---|---|
| **Sex** |  |
| Male | 7 (21.2) |
| Female | 25 (75.8) |
| Prefer not to answer | 1 (3.0) |
| **Age, years** |  |
| >25 to ≤35 | 4 (12.1) |
| >35 to ≤45 | 13 (39.4) |
| >45 to ≤55 | 11 (33.3) |
| >55 to ≤65 | 4 (12.1) |
| >65 | 1 (3.0) |
| **Years in practice** |  |
| 1 to <5 | 2 (6.1) |
| ≥5 to ≤10 | 5 (15.2) |
| >10 to ≤15 | 6 (18.2) |
| >15 to ≤20 | 6 (18.2) |
| ≥20 | 14 (42.4) |
| **Educational attainment** |  |
| High school | 2 (6.1) |
| Associate degree | 9 (27.3) |
| Bachelor’s degree | 13 (39.4) |
| Master’s degree | 8 (24.2) |
| PhD | 1 (3.0) |
| **Work Environment** |  |
| Hospital | 13 (39.4) |
| Academic | 10 (30.3) |
| Office | 9 (27.3) |
| Industry | 1 (3.0) |
| **Dominant scanning arm** |  |
| Right | 28 (84.8) |
| Ambidextrous | 4 (12.1) |
| Declined to answer | 1 (3.0) |

*Note*. N = number; PhD = Doctor of Philosophy; % = percentage.

### Effect of AI Assistance on Examination Time

AI-assisted scanning was associated with significantly shorter examination times compared with manual scanning, as shown in **Figure 4**. Mean examination time was 2.23 ± 0.44 minutes shorter with AI assistance than with manual scanning (95% CI: −3.10, −1.36; *P* < 0.001). A significant effect for Period was observed, with a mean reduction of 2.04 ± 0.44 minutes (95% CI: −2.91, −1.17; *P* < 0.001) in the second session, consistent with a learning or practice effect. A small but statistically insignificant difference in average exam time between Sequences (0.34 ± 1.08 minutes; 95% CI: −1.77, 2.46; *P* = 0.752), indicating that performance was not significantly influenced by the order in which participants utilized the AI-assisted and manual conditions.

**Figure 4.**
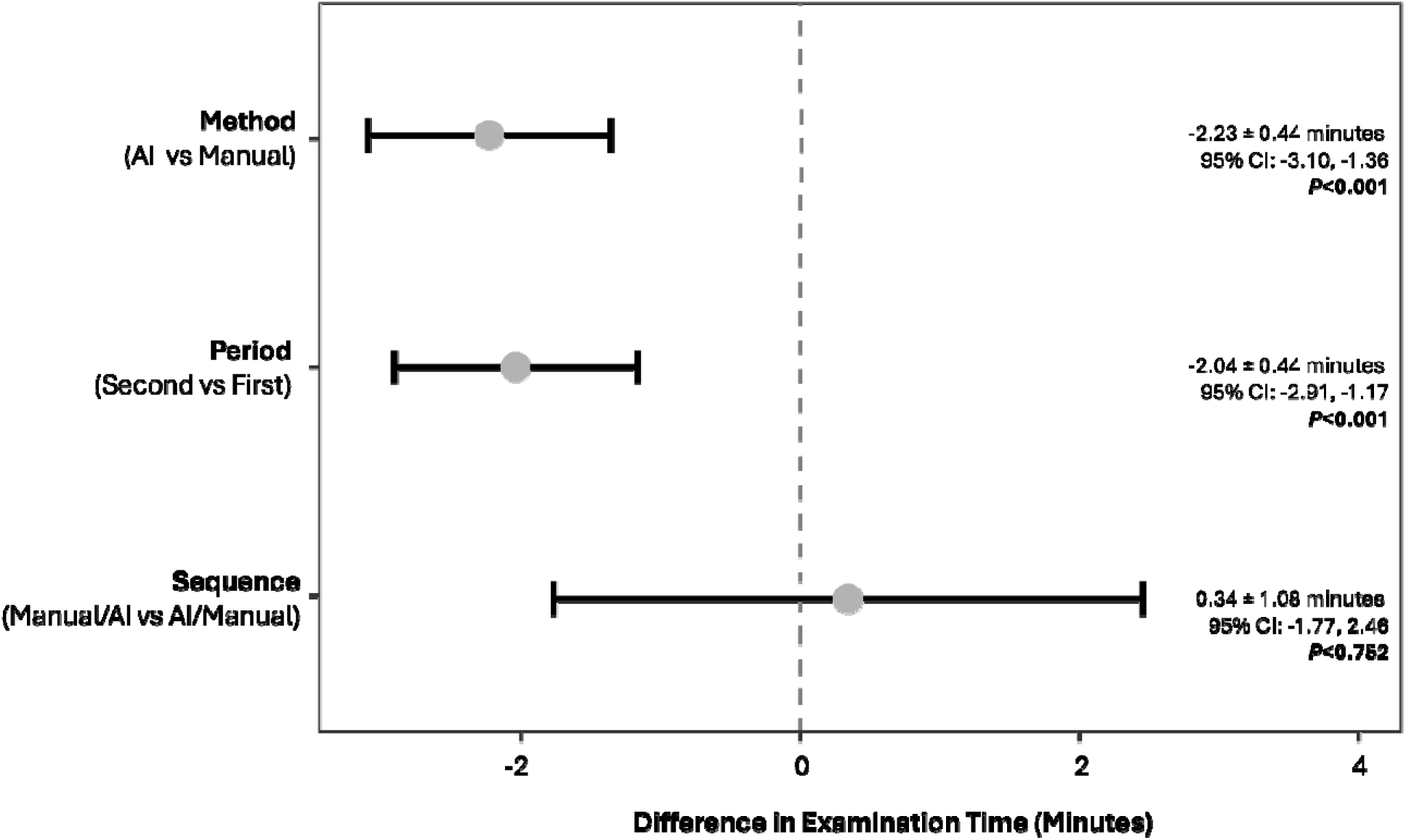
Difference in Examination Time by Method, Period, and Sequence. Mixed-effects model estimates with 95% confidence intervals.

Linear mixed-model estimated mean examination times by Method and Period are shown in **Figure 5**. Exam times improved for both Methods as participants became more familiar with the system after Period 1, however AI-assisted scanning was significantly faster than manual for both Periods.

**Figure 5.**
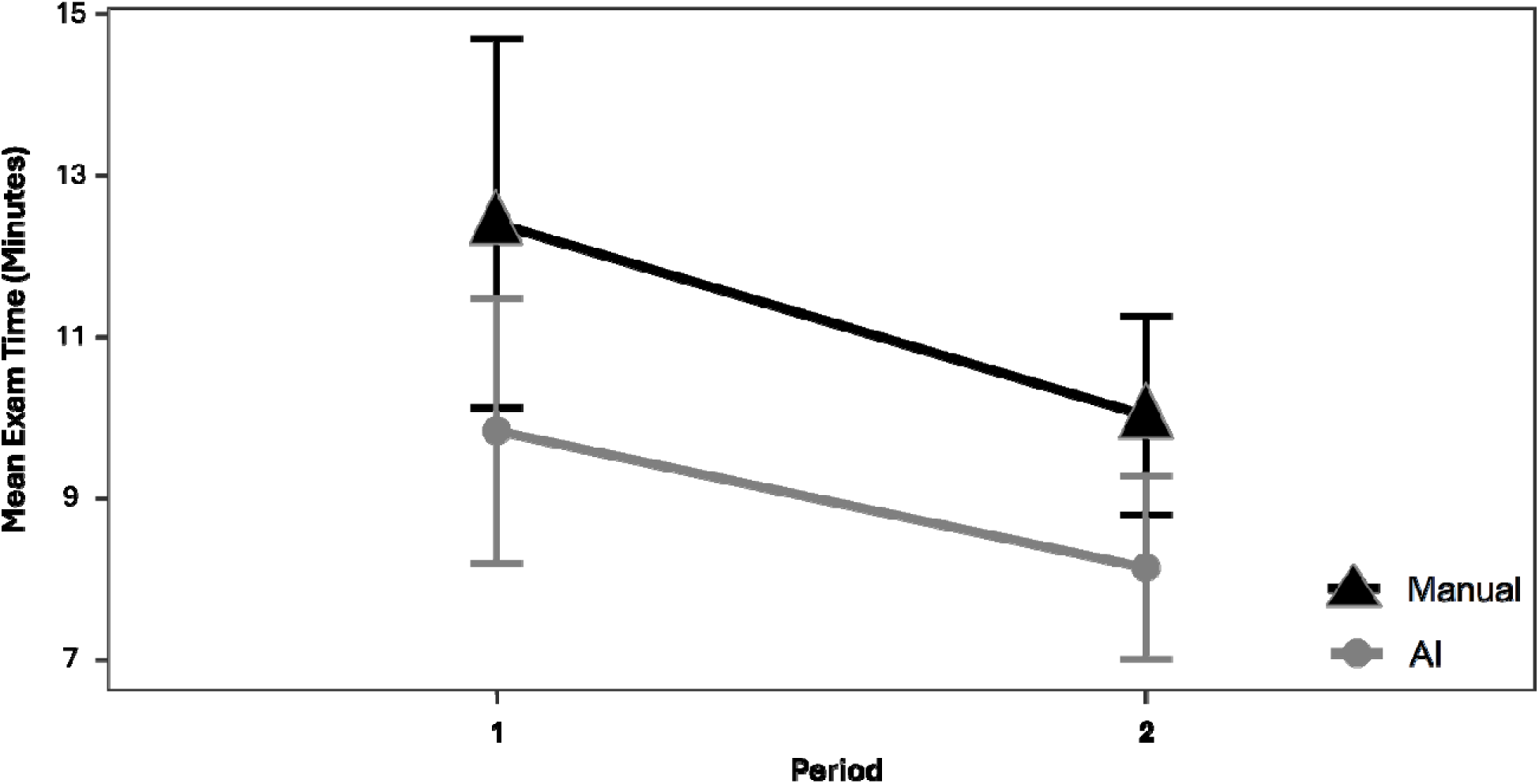
Mean Examination Time by Method and Period. Linear mixed model estimated means.

### Effect of AI Assistance on Ergonomic Burden

#### Distance Traveled (Non-Scanning Hand)

As summarized in **Figure 6**, AI-assisted scanning was associated with a lower ergonomic burden versus manual scanning. In a subset analysis (n = 17) utilizing a linear mixed-effects model, AI assistance was found to reduce distance traveled by the non-scanning hand by an estimated 22.91 meters compared to manual examinations (22.18 meters vs 45.09 meters; *P* < 0.001). A modest Period effect was also observed, consistent with a learning or familiarization effect. Of note, the effect of Sequence on distance traveled was minimal (see **Figure 6**) suggesting that order did not significantly influence the outcome and that carryover effects did not have an impact on the observed results.

**Figure 6.**
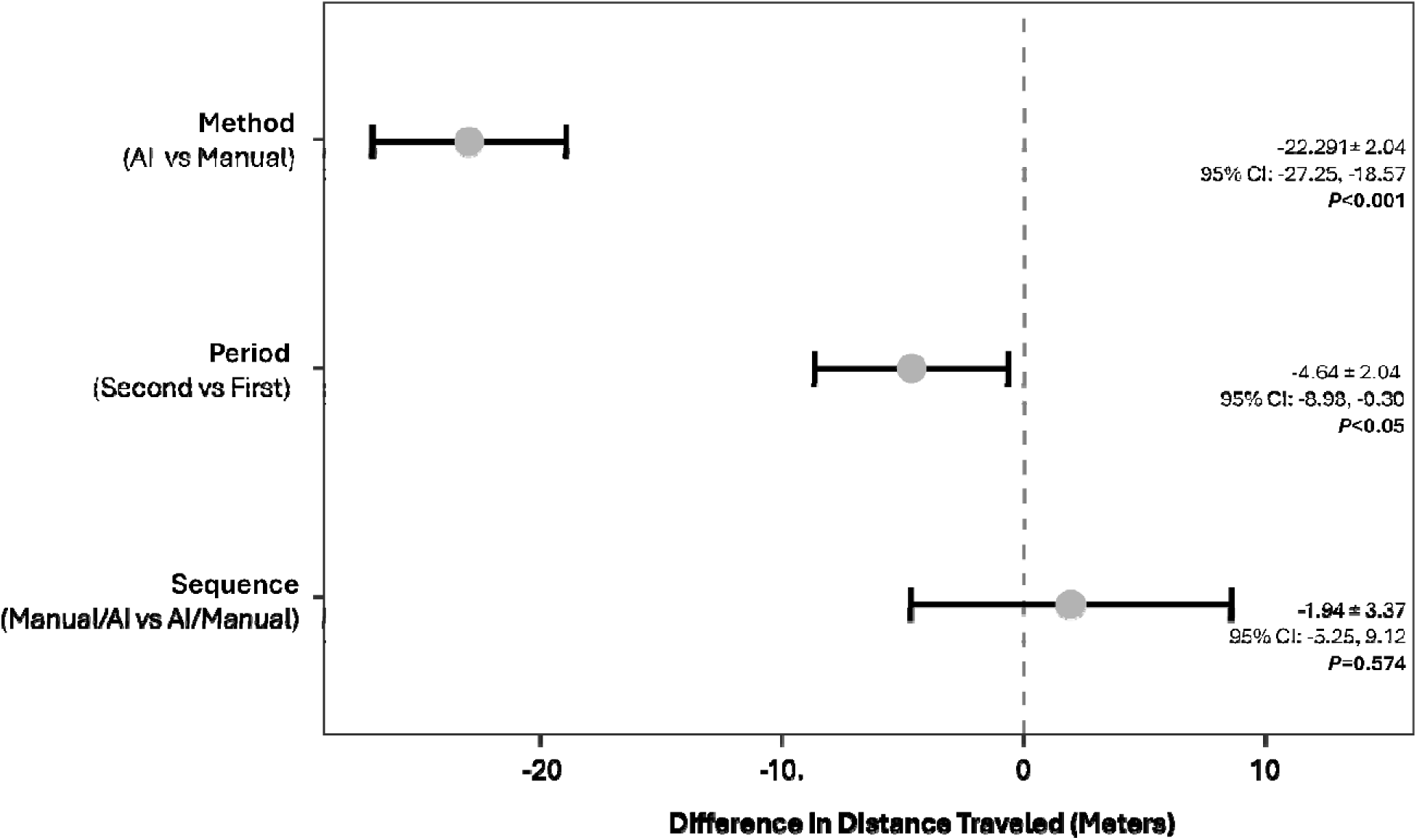
Difference in Distance Traveled by Non-Scanning Hand by Method, Period, and Sequence.

In addition to the overall reduction in distance traveled by participants’ non-scanning hands, observations of the motion paths with and without AI assistance suggest additional ergonomic benefit. As shown in **Figure 7**—which depicts the motion paths of a single participant during manual and AI-assisted scans—AI assistance was associated with a higher concentration of motion within the neutral ergonomic zone, with fewer reaches into the secondary and tertiary zones.

**Figure 7.**
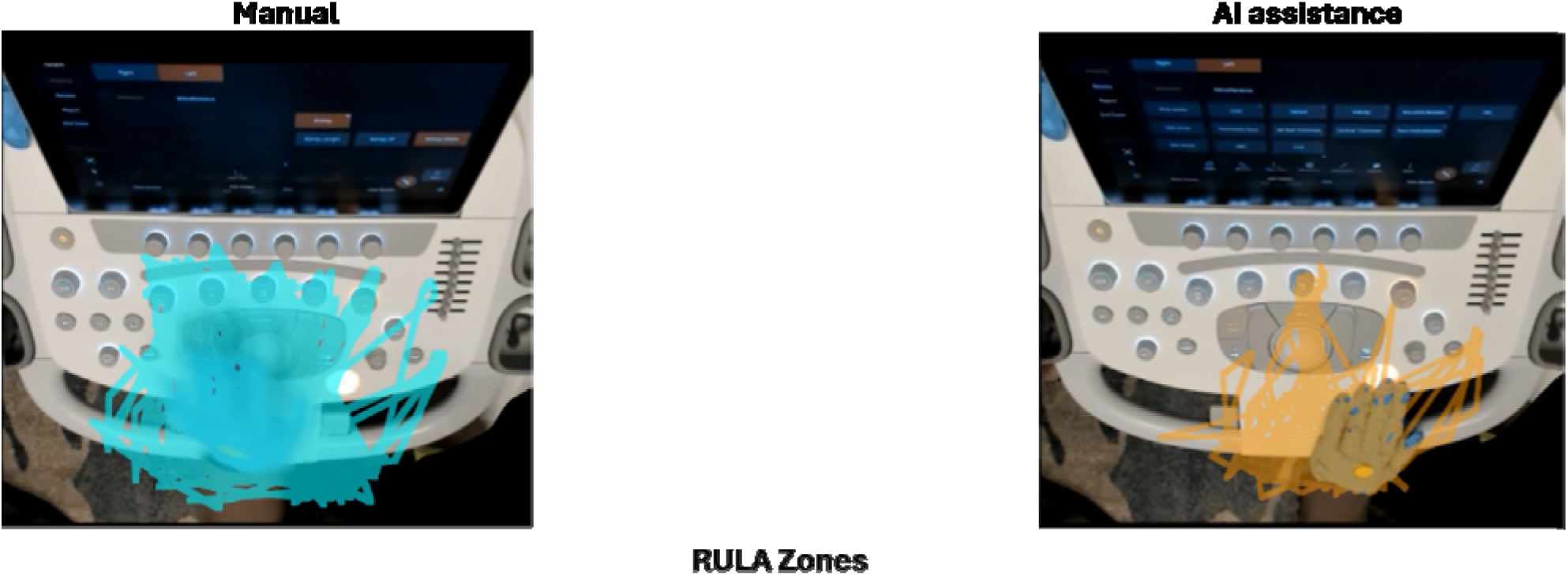
Motion Paths (Non-Scanning Hand) Without and With AI Assistance. Motion paths of an individual participant’s non-scanning hand during manual and AI-assisted image acquisition, illustrating both less movement and a higher concentration of movements in the neutral ergonomic reach zone. NOTE: lines represent movement of the tracked point on the dorsum of the middle finger; reach of the fingertip is a finger-length higher on controls. For full video of each scan, see **Supplemental Videos 1** (Manual) and **2** (AI-Assisted).

#### Interactions with System Inputs

**Figure 8** summarizes the estimated marginal means for overall click count and for each input type during manual and AI-assisted examinations. AI assistance was associated with a 36% reduction in the total number of clicks (*P* < 0.001), with differing levels of reduction across input types. Of note, touchscreen interactions—which are primarily in the secondary and tertiary ergonomic zones—were reduced by 41.5% with AI assistance. Neither Period nor Sequence significantly affected click counts.

**Figure 8.**
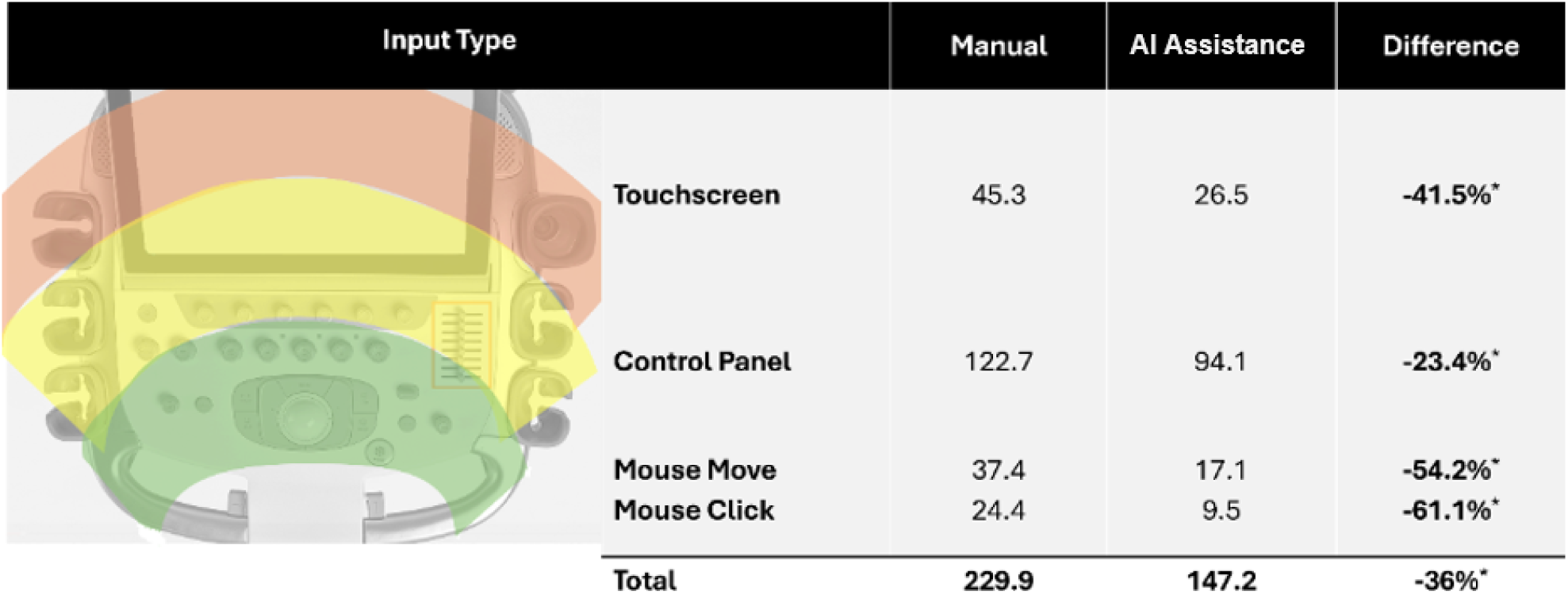
Click counts without and with AI assistance. Touchscreen = press of touchscreen and soft key knobs below touchscreen; Control Panel= press of controls on control panel; Mouse Move = movement of trackball; Mouse Click = press of either left or right mouse buttons.

### Effect of AI Assistance on Measurement Acceptance and Inter-User Variability of Offline Measures

In the offline measurement exercise, one participant completed only the Manual measurement portion of the task and was excluded from the analysis, resulting in 32 evaluable participants. Evaluable participants accepted 362 of 384 AI-generated measurements (94.3%) without modification. Nine measurements (2.3%) required a minor adjustment, and 13 measurements (3.4%) required a major adjustment. Inter-user variability as measured by the mean coefficient of variation was markedly lower for AI-assisted measurements: 0.019 (range: 0.000-0.091) versus 0.072 (range: 0.014-0.188) for manual measurements, representing a 74% reduction in variability.

### Qualitative Findings—Post-Study Survey Responses

Participant responses to the post-study survey are summarized in **Table 3**. Prior to the study, 41% of participants would have rated the benefits of AI assistance as high. After completing the study, 100% described AI assistance as having moderate (41%) or high (59%) benefit.

**Table 3.** Post-Survey Results (N = 33).

| Question | Participant Responses |
|---|---|
| Before this study, how would you have rated the potential benefits of AI in sonography? | • High Benefit 41.%<br>• Mod Benefit 26%<br>• Neutral 24%<br>• Low Benefit 6%<br>• Not Considered 3% |
| After this study, how would you rate the potential benefits of AI in sonography? | • High Benefit 59%<br>• Mod Benefit 41%<br>• No Benefit 0% |
| How would you describe your level of experience using AI-assisted tools in other healthcare settings? | • Extensive 6%<br>• Moderate 32%<br>• Limited 47%<br>• None 15% |
| Compared to other AI tools used, how effectively did this AI feature support your workflow? | • Sig More Effective 15%<br>• More Effective 55%<br>• Equal/Comparable 18%<br>• Did not Answer 12% |
| Based on your experience today, how do you see AI Abdomen impacting the overall workflow efficiency for routine abdominal exams? | • Significant Improved 24%<br>• Improved 56%<br>• No Change 3%<br>• Reduced 9%<br>• Significant Reduced 9% |
| With AI Abdomen enabled, how would you rate the accuracy of the AI view recognition and labeling? | • Very Accurate 24%<br>• Somewhat Accurate 18%<br>• Accurate 59% |
| Based on your experience today, how would you rate the accuracy of the AI measurements? | • Very Accurate 29%<br>• Somewhat Accurate 21%<br>• Accurate 50% |
| Based on your experience, how do you see AI Abdomen impacting the overall exam time of routine abdominal exams? | • Sig Reduce Time 32%<br>• Reduce Time 62%<br>• No Change 3%<br>• Increase Exam Time 3% |
| Based on your experience today, how do you see AI Abdomen affecting the amount of hand movement for routine abdominal exams? | • Significant Reduction 35%<br>• Reduction 53%<br>• No Change 6%<br>• Increased Movement 6% |
| Based on your experience today, how do you see AI Abdomen affecting consistency across different users? | • Significantly Improved 26%<br>• Improved 71%<br>• No Change 3% |

*Note*. AI = artificial intelligence.

Eighty five percent of participants had at least some experience using AI-assisted tools in other healthcare settings. When asked how the study’s AI assistance compared to other tools in supporting their workflow, 68% of participants indicated it was more effective. Eighty percent also indicated they believed it would improve (56%) or significantly improve (24%) overall workflow efficiency for routine abdominal examinations, and 94% of participants predicted that use of AI assistance would reduce (62%) or significantly reduce (32%) overall exam time.

All participants found AI view recognition, labeling, and measurement to be accurate, with approximately 25% classifying these functions as “very accurate.” In addition, 97% of participants indicated that based on their experience in the study, they believe AI assistance would improve (71%) or significantly improve (26%) the consistency of abdominal ultrasound examinations across different users.

## DISCUSSION

Enhancing scan efficiency is a critical goal in sonography, both from the perspective of increasing throughput to match demand and for reducing sonographer physiologic stress and workload. In this randomized crossover study, AI-assisted scanning using the ACUSON Sequoia ultrasound system with AI Abdomen led to statistically significant and potentially meaningful reductions in the time required to complete a standardized abdominal ultrasound examination and improved the ergonomics of movement by participants’ non-scanning hands. These data suggest that AI assistance may reduce repetitive stress and time spent in awkward positions that can contribute to the development of WRMSDs.

In addition to shortening exam times by an average of 2.23 minutes, AI assistance reduced the average distance traveled by the non-scanning hand by 22.91 meters. This represents a 49.2% reduction in overall hand movement required to operate the system console, potentially reducing ergonomic strain on the non-scanning shoulder and arm. Of note, there was no evidence of sequence or carryover effects for the reductions in time and distance, supporting the validity of the crossover design and strengthening confidence that the observed effects are true improvements rather than artifacts of treatment order or residual influence from the prior study period.

Analysis of hand interactions with specific system inputs provides further insights into the ergonomic benefits of reduced hand movement. The trackball on the ACUSON Sequoia control panel is positioned so the user’s hand, elbow, and shoulder joints are in a neutral position during typical use. To reach the touchscreen and other inputs on the control panel, users must reach forward out of the neutral zone into secondary and tertiary ergonomic zones, as illustrated in **Figure 3**. In this study, AI assistance was associated with a 41.5% reduction in touchscreen interactions and trackball moves and mouse clicks were reduced by 54.2% and 61.1%, respectively, indicating fewer repetitive motions that can negatively impact the wrist.

Importantly, these gains were achieved among sonographers without prior experience with AI Abdomen, suggesting that AI assistance can be beneficial to practicing sonographers, regardless of prior AI experience.

The agreement and inter-user consistency of AI-assisted measurements were demonstrated during an offline measurement exercise in which 94.3% of AI measurements were accepted without modification and AI assistance reduced inter-user variability by 74%. In addition, 100% of participants found the AI Abdomen view recognition, anatomical labels, and measurements to be accurate. This perceived precision is a critical factor for reproducibility and standardization of abdominal examinations across operators and suggests that the improvements in scan time seen with AI assistance were not achieved at the cost of accuracy and reproducibility.

Post-study survey responses indicate that the study altered participants’ perception of the benefits of AI: the proportion of participants who considered AI tools to be beneficial rose from 67% prior to the study to 100% after study completion. Survey responses also confirmed the ease and functionality of AI-assisted technology, with participants consistently reporting ease of use, improved workflow efficiency, and greater measurement consistency.

This study has several limitations. Because a single ultrasound system model was used, findings are specific to the ACUSON Sequoia system with AI Abdomen enabled. Although efforts were made to ensure a balance of participants with varied experience, credentials, education, and age, there is probably some degree of selection bias, as sonographers who volunteered may be different from those in the general population. The imaging models used in this study were of normal body habitus; thus, the study provides no information on the utility of AI-assisted scans in more challenging patient populations in clinical settings. While the reduction in distance traveled during scans was substantial and may potentially reduce WRMSDs, the direct impact on work-related injury was not studied.

Overall, this study provides evidence that AI-assisted technology can improve scanning efficiency and reduce ergonomic burden during routine abdominal ultrasound. Time gained because of shorter scan times could be used in a number of ways, including potentially increasing patient throughput. Additional studies on the use of AI Abdomen in clinical settings can further clarify the real-world impact of this technology and its potential for supporting efficient, ergonomically sustainable workflows.

## Data Availability

All data produced in the present work are contained in the manuscript

## FUNDING

This study was sponsored and fully funded by Siemens Healthineers. Siemens Healthineers provided financial support for institutional review board approval, investigator compensation, participant compensation, model compensation, and study operations. AIUM recruited and coordinated study participants, with supplemental onsite participant recruitment by AIUM staff. Siemens Healthineers participated in the study design, study conduct, data collection, analysis, interpretation, and assisted with manuscript preparation.

## CONFLICTS OF INTEREST

**RGB**: Siemens Healthineers – Research grants, Speakers Bureau; Philips Ultrasound - Research grants, Speakers Bureau; Mindray - Research grants, Speakers Bureau; Canon Medical - Research grants, Speakers Bureau; Samsung Ultrasound - Research grants, Speakers Bureau; Fujifilm - Research grants, Speakers Bureau; Bracco Diagnostics – Speakers Bureau, Advisory Panel; Olea Medical – consultant; Treasurer – International Contrast Ultrasound Society (ICUS) – nonpaying position; Secretary General – International Ultrasound Medical Ultrasound Alliance Committee – (IUMAC) nonpaying position; Editor-in Chief Journal of Ultrasound in Medicine (JUM) official journal of American Institute of Ultrasound in Medicine.

**JG**: Canon Medical Systems - Research grants, Speaker honorarium

**AL**: received speaker honorariums from Canon Medical systems, Siemens Healthineers, Samsung Ultrasound, GE Healthcare, Fujifilm and Bracco. He has received research grants from Philips Ultrasound and GE Healthcare, and equipment grants from Canon Medical Systems, Samsung Ultrasound, GE Healthcare and Siemens Healthineers. He receives royalties from Wiley Publishers.

**BN**: None

## ACKNOWLEDGMENTS

Editorial assistance was provided by Catherine Grillo, MS, of BioScience Communications, and funded by Siemens Healthineers. Siemens Healthineers supported this research study.

## Notes

### Competing Interest Statement

rgb- Research grants - Siemens, Philips, Mindray, Samsung, Vinno, SonoScape,Canon, GE. Speakers Bureau- Philips, Vinno, SonoScape, Canon, Bracco Diagnostics. Advisory panel - Bracco Diagnostics
AL -Siemens Healthineers Equipment grant and speakers bureau
JG -Siemens medical Solutions grant and speakers bureau
BN - nothing to disclose

### Author Declarations

WCG Investigational Review Board #20251081. All participants and sonographers signed informed consent documents.

